# Health care workers’ level of adherence to infection prevention and control and its impact on puerperal and neonatal sepsis among delivering women and neonates in Central Tanzania: A protocol for prospective cohort study

**DOI:** 10.1101/2022.12.18.22283631

**Authors:** Laura Edward Marandu, Golden Masika

## Abstract

**Introduction:** Delivering women and neonates are at a great risk of acquiring infections due to lack of adherence to infection prevention and control (IPC) and low level of immunity and extended exposure to care procedures that can lead to infections. This prospective study aims to assess the level of adherence to IPC among health care workers and its impact on puerperal and neonatal sepsis in Dodoma region.

**Methods and Analysis:** Level of adherence to IPC is examined cross-sectionally among health care workers (HCWs’) in contact with delivering women and their neonates. Prospective cohort approach is used to assess level of exposure of 294 delivering women and their neonates to poor hygienic practices of HCW through observation checklist. Outcomes including incidence of puerperal and neonatal sepsis are evaluated clinically two days later before discharge. Laboratory culture and sensitivity confirmatory tests of blood samples are done on positive cases. Data analysis for level of adherence to IPC practices, incidence of puerperal and neonatal sepsis as well as relative risk among the exposed women and neonates will be performed.

**Ethics and Dissemination:** The University of Dodoma Research Ethics Committee (UDOM-REC) approved this study (Ref No. MA.84/261/’A’/25). Findings of this study will be published in international peer reviewed journals, disseminated in international conferences, to the participating hospitals, the University of Dodoma and the Tanzanian Ministry of Health for informing practice and policy.

**Strengths and limitations of this study:**

- By capturing data from both health care workers and patients using mixed methods approach, the study can examine the impact of health care worker’s infection prevention, and control practice on patient’s outcomes at the same time.
- Compared to other studies, this study objectively tracks and quantify the risk of infection from all possible points of contact between a health care worker and a client.
- The study can enable early identification of puerperal or neonatal infection and initiate treatment before discharge Exposure to pre-admission infection long incubation period and manifest at the hospital post-delivery may be difficult to capture and may confound the results without culture and sensitivity.
- The post-delivery women and their neonates will be followed for two days only to abide to hospital discharge protocol; this will limit the data to early onset of sepsis only.

## Background and rationale

Globally infection prevention and control (IPC) practices and maternal outcomes remain a great challenge especially in the middle and low income countries due to low level of technology, understaffing and limited medical supplies[1]. Worldwide 15 out of 100 patients/clients who receive care in health care facilities develop infections which are related to poor adherences to infection prevention and control standards including maternal and neonatal infections[2]. Maternal infections before or during child birth contribute to approximately one million newborn death annually. These deaths are contributed by postpartum infection, pre-existing maternal conditions such as malnutrition, diabetes, obesity, anemia, bacterial vaginitis and group B streptococcus infections; spontaneous or providers initiation conditions during labor and child birth such as premature rupture of membranes, multiple vaginal examinations, manual removal of placenta and caesarean section[3].

In reaching Alma Ata declaration and ensuring health for all, health care standards including adherence to IPC standard must be complied in achieving better outcome and minimizing the risk of puerperal and neonatal sepsis[4]. Adherence to IPC standards during labor and child birth can reduce the risk of acquiring puerperal and neonatal sepsis. On the contrary, failure to adhere to IPC standards in labor wards can leads to health care associated infections (HCAIs) to delivering women, newborns and health care workers[5]. This in turn, may results into morbidity and mortality of patients, increased treatment costs for both patients and health care system due to increases length of hospital stay and additional investigations ordered[6].

In the African settings where financial and/or health care resources to combat or manage HCAIs to delivering women and their neonates are scarce, adherence to IPC standards is vital in prevention of such infections from occurring. Thus, practice related factors such as hand washing using soap and running water before and after any procedure, appropriate use of personal protective equipment (PPE), frequency of per vaginal examinations (PVE), control for duration of labor and rupture of membranes, use of sterile, clean or highly disinfected equipment, environmental cleaning, waste and linen management that have shown to reduce the risk mortality related to puerperal and neonatal sepsis must be considered in prevention of infection in the health care facilities[7,8]. In addition, service related factors such as number of Antenatal clinic (ANC) visits, minimizing delivery by cesarean section (C/S), presence of infection control committees and routine maternal audits in the health facilities have also shown to reduce the risk mortality related to puerperal and neonatal sepsis[8].

Nevertheless, researchers have identified a number of factors that heighten the rate of puerperal and neonatal sepsis. Such situations include inadequate compliance with IPC standards during cesarean section[9]. Others include poor personal hygiene, rural residence, lack of proper equipment sterilization, postpartum hemorrhage, anemia, prolonged labor and bacterial infection to be directly associated with puerperal sepsis[10–12]. In addition, inadequate training on IPC and stock out of basic consumables and equipment’s such as surgical gloves and elbow tap, have shown to contribute to poor adherence to IPC standards and increase the risk of infection to mothers and their newborns especially in the sub-Saharan Africa[13].

The relationship of these variables and the risk of puerperal and neonatal sepsis can be best explained by the Epidemiological triad theory that explain about development of a disease as a results interaction between the agent and the susceptible host in an environment that supports transmission of the agent from a source to the host.[14,15]. Altered balance and interactions of the three components must occur such that the susceptibility of the host and presence of an environment that exposes the host to an agent are key to rendering infection/disease to occur[16]. In the context of obstetric care, delivering women and neonates are susceptible to various nosocomial infections and from the community settings due to suppression immunity during pregnancy[17], whereas neonates have immature immune system to fight infections in the microbial-rich environments[18]. Figure 1 illustrates the interaction of the variables of the Epidemiological triad as explained by Johnson-Walker & Kaneene [14]

**Figure 1.**
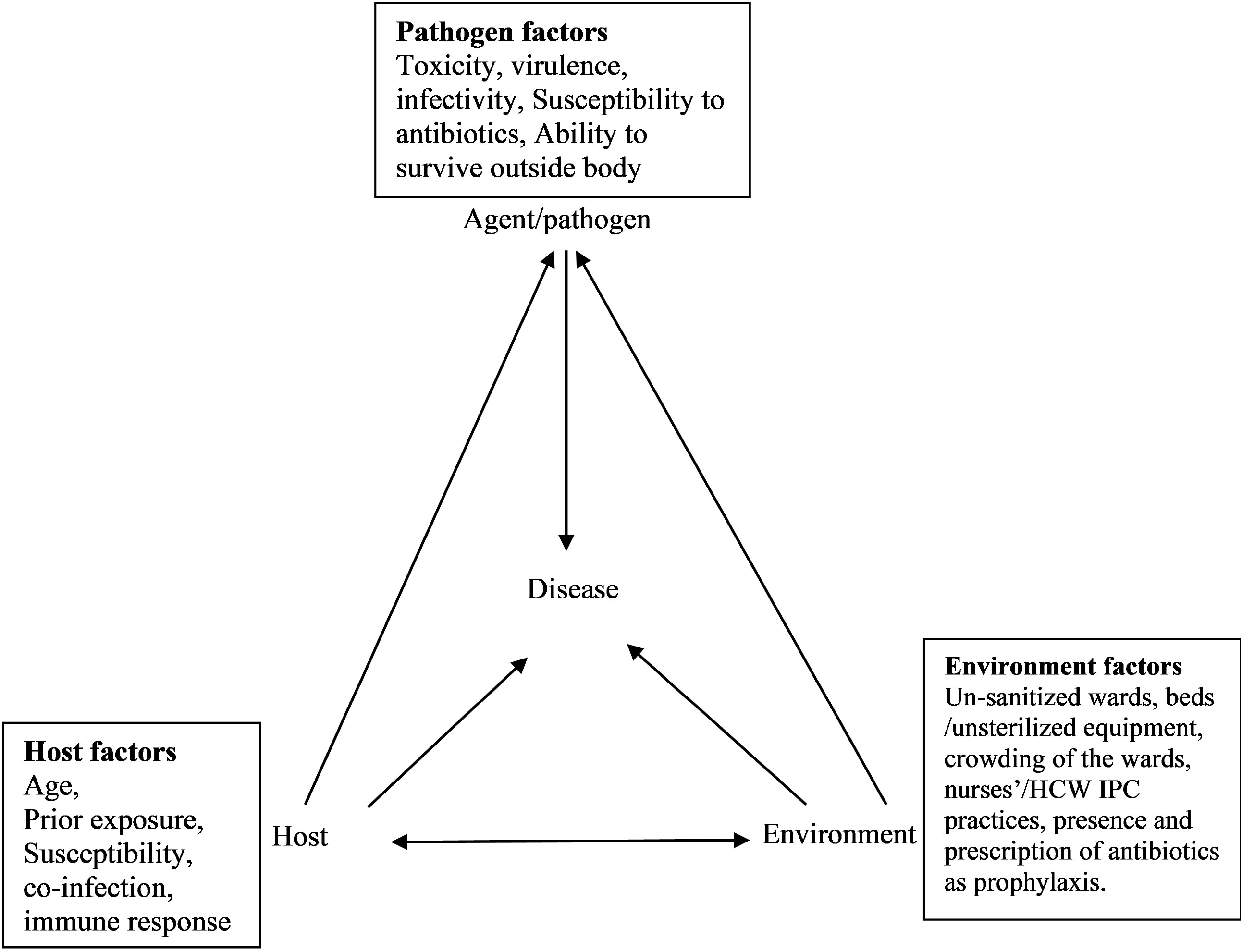
Interaction of the variables of the Epidemiological triad as adapted from Johnson-Walker & Kaneene,[14].

In Tanzania, adherence to IPC standards in obstetric care has additional variables that contribute to the complexity of the issue. Example, a Star Rating Assessment (SRA) in Tanzania reveals that with only 31% prevalence of adherence to IPC standards, the private health care facilities have better adherence compared to the public health care facilities[19]. Also, tertiary and secondary health facilities have better adherence to IPC standards compared to primary health care facilities[19]. Despite of literature providing a clear link between poor adherence to IPC standards in the context of obstetric care and the risk of puerperal and neonatal sepsis, it is to the best of our knowledge that the amount of risk attributed by the exposure to the incidences of non-adherence to IPC standards and the magnitude of risk to puerperal and neonatal sepsis among delivering women and their neonates in Tanzania has never been studied. Therefore, this study uses a descriptive cross-sectional approach to examine the pattern of HCWs’ adherence to IPC standards, and a prospective cohort design to examine the risk of exposure to HCWs non-adherence to IPC standards on puerperal and neonatal sepsis among delivering women and their neonates in Dodoma.

### Aim

This prospective study aims to assess the level of adherence to IPC among health care workers and its impact on puerperal and neonatal sepsis in Dodoma region. The specific objectives are;

1. to determine level of adherence to IPC standards in labor and postnatal wards among HCWs’ in Dodoma region,
2. to determine the incidence of puerperal sepsis from day of admission to day of discharge among postnatal women,
3. to determine the incidence of early neonatal sepsis among neonates,
4. to examine the risk of puerperal sepsis among post-delivery women attributed by exposure to HCWs’ poor IPC practices.
5. to examine the risk of neonatal sepsis among neonates attributed by exposure to HCWs’ poor IPC practices
6. to determine other risk factors independently associated with development of puerperal and neonatal sepsis.

## Methods and analysis

### Study design

This study adopts a cross sectional approach for the first objective to examine the level of adherence to IPC standards in labor and postnatal wards among HCWs; and a prospective cohort design[20,21] is adopted for examining the incidence of puerperal and neonatal sepsis and the amount of risk in developing puerperal and neonatal sepsis. After the baseline demographic data are taken a woman and her neonate begin to be followed up on all the points of possible exposures to infection immediately after admission to labor ward and post-natal ward. Thus, observation checklist is used to assess practice of HCWs’ and score for practice events that IPC standards are not followed. This will enable gauging the level of adherence to IPC standards required at the labor and postnatal wards as prescribed in the National guidelines. Another observational checklist is used at baseline to score for all events that a woman and her neonate are exposed to practice incidents where IPC standards are not followed. Each of those events are scored and ultimately be used to compute the magnitude of exposure and the incidence rates and relative risk of puerperal and neonatal sepsis across different levels of exposure to poor IPC practices. The recruitment of participants began in April 2022 and is expected to end in December 2022

### Study settings

The study is being conducted in Dodoma region involving three hospitals. Dodoma Regional Referral Hospitals (DRRH), St. Gemma Designated District Hospital (DDH), and Makole Municipal Health Centre; all situated in Dodoma city. Dodoma Region is a capital city of Tanzania. Projection data of 2019 from the National Bureau of Statistics show that Dodoma has a total population of 2,492,989 with annual growth rate of 2.1.[22]. Found in central Tanzania, Dodoma has 7 districts in an area of 41,311 square kilometers[22]. There are a total of 223 dispensaries and 32 health centres that make the referral sources for the more complex obstetric cases to the DRRH, whereas also complex cases from the St. Gemma DDH and Makole Municipal Health Centre are also referred at the DRRH.[22].

Owing the fact that Dodoma city is fast growing as a result of high number of immigrants following shift of Government activities (Ministries) from Dar es Salaam to Dodoma, there has been a tremendous increase in the demand for health care services including obstetric care services[22]. Even with this demand, Dodoma is one of the regions that perform poorly in star rating as evaluated by the Ministry of Health where IPC adherence was one of the domains assessed[23]. Due to these alarming data, it is justifiable to conduct this study in Dodoma and provide empirical evidence for future directions towards interventions to curb this problem.

### Study population

This study involves health care workers, delivering women and their neonates as study populations. Health care workers in the labor wards and post-natal wards are targeted for assessing the level of adherence to IPC standards related to their practices. Their practices determine exposure to infectious agents among delivering women and neonates; thus, they are studied for adherence to IPC standards during interaction with delivering women. Women and neonates are the target populations who are subject to the outcomes of adherence/non-adherence to IPC standards of health care workers’ practices. Delivering women are studied starting from baseline, on their level of exposure to health care workers’ poor IPC practices from the day of admission to labor ward until postnatal discharge. The neonates are studied immediately after is born until discharge.

### Inclusion and exclusion Criteria

#### Inclusion Criteria

*For HCWs’*

HCWs who work in the labour ward and postnatal ward are included

*For Women*

Women who deliver by spontaneous vaginal delivery (SVD) at a respective research site during the time of research are included

*For Neonates*

All neonates delivered by women who are studied, also are included.

#### Exclusion Criteria

Health care providers who meet the inclusion criteria but refuse to consent are excluded. Delivering women who present with conditions/complications such as preterm baby, pre-rupture of membrane, signs of existing infection, postpartum hemorrhage (PPH) and fever are excluded as they can bias the evaluation of outcomes. Women who delivered by SVD but before arriving (Birth Before Arrival) to the facility of a respective study area are also excluded. A neonate is excluded from study if seriously sick of other conditions not related to puerperal sepsis.

#### Sample size determination

The sample size for cohort study is calculated based on the following formula as adopted from (Kelsey 1996) taking assumptions of one cohort group where some of the subjects will be exposed to poor IPC practices for HCWs’ and others not exposed.

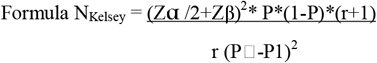

Where:

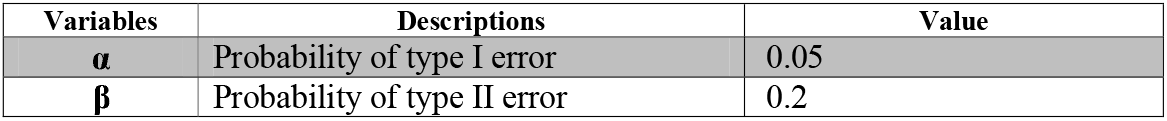
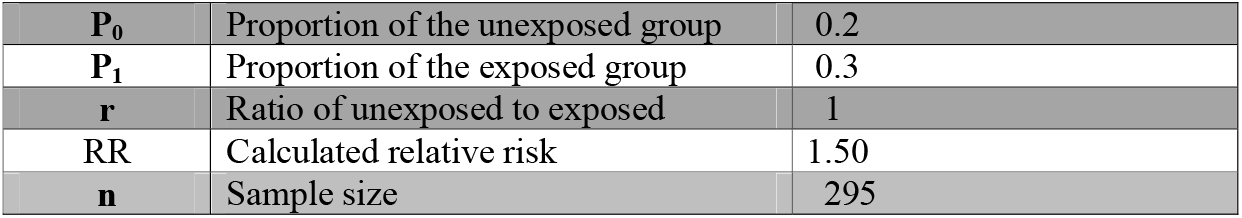

Based on Kelsey’s formula, the sample size will be 295 delivering women. Every neonate of a woman recruited in the study will be also recruited. Thus, if all women will have single tone pregnancy, 295 neonates will be included in the study.

As HCWs working in the labor and postnatal wards are few, the sample size calculation for this group is not warranted. Instead, all available HCWs were asked to participate.

#### Sampling procedure

The sample is being drawn from 3 health care facilities purposively selected because they have large volume of deliveries per day to represent three levels; regional referral hospital, a district hospital and a health centre. Thus, the health care facilities include Dodoma RRH, St Gemma DDH and Makole Municipal Health Center.

Proportional sampling is used where a stratum sample size is allocated in proportion to population size within stratum. i.e.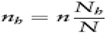. This procedure is done to determine the proportional samples for women and then their newborns. Women are being recruited following a systematic random sampling procedure until a daily sample size is reached. As for Dodoma RRH and Makole HC both with a maximum number of 20 SVD per day the daily sample of 10 is enrolled by recruiting every second mother giving birth in the delivering room whereas in St. Gemma DDH with a maximum of 10 deliveries per day, 5 women are recruited using the same systematic random sampling procedure. Neonates of women included are also included in the study after they are born. As for HCWs, all available at the labor ward and postnatal ward who interact with delivering and postnatal women at the facilities under study were invited to participate at the beginning of the study.

#### Variables and measurements Outcome Variables

Two outcome variables are considered in this study. We measure the incidence of puerperal sepsis for post-delivery women and incidence of neonatal sepsis for neonates as outcome variables. The two outcome variables are measured as follows:

#### Puerperal Sepsis

Puerperal sepsis is evaluated using clinical signs and symptoms by the doctor. A woman is considered to have puerperal sepsis if found to have lower abdominal pain, abnormal vaginal discharge with a foul smell plus or minus fever >38°C. If the signs and symptoms persist after administration of antibiotics laboratory investigations including culture and sensitivity so as to isolate the specific microorganism responsible for the infection is performed. This diagnostic approach is a gold standard for diagnosis of puerperal sepsis in Tanzania and considered reliable for this study. During codding, the measure of puerperal sepsis is dichotomized into (i) puerperal sepsis, coded as 1 (ii) no puerperal sepsis, coded as 0.

#### Neonatal Sepsis

Neonatal sepsis is evaluated using clinical signs and symptoms by a doctor. A neonate is considered to have neonatal sepsis if he/she is found to have temperature >38°C, foul smelling cord stump, discharge from the cord, lethargic, jaundiced, and/or conscious/unconscious. If the signs and symptoms persist after administration of antibiotics laboratory investigations are done where blood culture and sensitivity is done to rule out any of the microbial agent from the child’s blood sample. This diagnostic approach is a gold standard for diagnosis of neonatal sepsis and is routinely used in the context of Tanzania. Same as for puerperal sepsis, the measure of neonatal sepsis is dichotomized into (i) neonatal sepsis, coded as 1 (ii) no neonatal sepsis, coded as 0.

## Predictor variables

### Baseline Demographic characteristics

For health care workers, demographic characteristics including age, sex, marital status, occupation, profession and working experience are collected. As for delivering women demographic characteristics collected at baseline include age, marital status, level of education, occupation and residence.

As for neonates, demographic data collected at baseline include neonate birth weight, feeding status, gestational age, Apgar score and sex.

### Level of adherence to HCWs’ hygienic practices

Level of adherence to IPC standards is measured using IPC observational checklist among HCWs; the observational IPC checklist was adopted from the standard Hospitals Assessment Tool on Standards-Based Management and Recognition (SBMR) for Improving IPC of the Ministry of Health (MoH). The checklist has 7 domains with total of 65 items which are measured, each item is scored as yes (1) or No (0). The highest score for all items is 65 and the lowest is 0 with higher score indicating adherence to IPC standards.

### Level of exposure to health care workers hygienic practices

Immediately after admission to labor ward, women are examined for signs of infection by history taking and observation to rule out any infection before inclusion in the study. Those who are free from infection are included and start follow up through observation for level of exposure to unhygienic practices of HCWs’. The observation checklist was developed by the research team comprising of 4 domains with 12 items. Observation points indicating a woman is subjected to infection are captured by the checklist as baseline exposure. These include the following domains (i) hygiene of the bed for a woman to transfer into; has 3 observation items, (ii) during per vaginal examinations; contains 4 items; (iii) during injections has 1 item, and (iv) during delivery of the placenta, has 4 items.

As for the neonate, exposure to hygienic practices of HCWs’ as a baseline measure is based on 5 domains with 22 items which are captured from checklist observation points that include (i) contact with a HCWs’ during delivery; 6 items, (ii) during resuscitation; 6 items, (iii) during comprehensive neonatal assessment; 4 items, (iv) immunization; 3 items, during skills training to a woman on how to properly breastfeed; 3 items. These items are formulated based summing up the activities of the routine labor ward and postnatal ward practices and directives of the obstetric care guidelines in the context of Tanzania.

Items in both woman’s and neonate’s checklists are scored as “Observed” and given a score of 1 when a HCWs’ shows a deviation from the recommended IPC standard, and scored as “Not observed” and given score of 0 when practices according to the recommended standard. The checklist for a woman therefore, have the score ranging from 0 to 12 whereas that of neonate score from 0 to 22. The higher the score the higher the level of exposure to unhygienic practice of HCWs’.

### Data collection procedure

#### Observations

After they consented to participate in this study, HCWs’ in each facility are being observed for their adherence to IPC standards during practice in labor and postnatal wards. Data are collected by the trained nurses who are staff in other health care facilities different from study sites. On the first day, observation of all HCWs’ practice was conducted and data were recorded. However, those data are assumed to have been influenced by Hawthorne effect and potentially lead to observation bias due to presence of the observer on the first instance when they are informed to being observed. Thus, these data will be excluded from analysis. Actual observation of practice and data collection began on the second day. Multiple events of the HCWs’ while attending patients are being observed and data are abstracted in the checklist.

The same nurses who collect data from the HCWs, also observe and abstract data for women’s and neonates’ exposure to poor IPC practices using a different checklist. Every encounter of a woman and an HCW is observed and recorded as to whether “observed” of “not observed” and given the score of 1 and 0 respectively according to items

#### Procedure for collection and transport of blood sample for culture

Trained nurses collect blood samples from mother and/or neonate who show signs of infection for investigation by observing IPC standards and standard operating procedure (SOPs) for blood sample collection and transportation. They use 1-2 mls in ethylenediaminetetraacetic acid tube (EDTA) tube collected by vacutainer needle and sent to the laboratory for Full Blood Picture (FBP) where processing in three- or five-parts differential machines is performed. They also collect 5 – 10mls 5-10mls of blood into two blood culture bottles (Aerobic and Anaerobic bottles) for culture and sensitivity as per hospital protocol and manufacturer’s instructions. The blood culture is incubated in 37CC for 7-14 days. Therefore, if significant growth is seen, isolation, identification, and drug sensitivity of microorganisms are examined.

#### Data management and analysis plan

Data collected are checked for quality and kept with confidentiality per the University of Dodoma’s (UDOM) policies. They are kept safe under the computer with password, accessed only accessed by the researchers. After the study is completed and the findings are published, the chiliasts and questionnaires with individual data will be destroyed and the electronic data set will be kept by the researchers per UDOM’s policies.

After all the data are collected, analysis will be performed using Statistical Package for Social Sciences (SPSS) version 25.0. Descriptive analyses will be used to summarize the demographic characteristics of HCWs, women and neonates. Pattern of HCWs’ adherence to IPC standards in their practice is set to be analyzed using descriptive statistics of mean/median and ANOVA to compare the overall mean score per IPC domains for each of the three hospitals; and logistic regression. where the association between adherence and demographic characteristics will be examined. The incidence of puerperal sepsis and neonatal sepsis will be computed using number of cases of puerperal sepsis per person time obtained from the number of people followed up and the summation of time each one contributed before developing puerperal sepsis. Same approach will be used for calculation of incidence for neonatal sepsis. As for risk of exposure to HCWs’ poor IPC practices and development of puerperal sepsis, rating categories defined by the Ministry of Health as determined by adherence scores will be used to determine the threshold for high exposure and low exposure groups. Then, the risk ratio will be calculated to compare risk of developing puerperal sepsis among those with high exposure to HCWs’ poor IPC practices and those with low risk. Similar approach will be used to calculate the risk ratio for developing neonatal sepsis. Otherwise, to examine the influence of various factors including adherence to IPC practice on development of puerperal and neonatal sepsis and control for potential confounders, an overall multivariate logistic regression analysis will be performed. Results will be considered statistically significant at p < 0.05 using 2 tailed tests and estimates will be presented with their 95% confidence intervals.

#### Ethics and Dissemination

This study obtained an ethical approval by the University of Dodoma Reserch Ethics Committee. Permission to conduct this study has been sought from the President’s Office, Regional Administration and Local Government Authority (PO-RALG) and from the Dodoma Regional Administrative Secretary (RAS). Each participant who is being recruited in this study signs the informed consent and those who cannot read and write receive oral information about the study and thumb print on the consent form. As for the neonates, assent by the parent is obtained. To enhance anonymity and confidentiality of participant’s information each questionnaires and checklists are identified with codes and numbers instead of participant’s names. All data are kept secured by the principal investigator in a locked cupboard until allowed time for discarding per UDOM’s policies.

The findings of this study will be disseminated to the following avenues. First the findings will be disseminated to the participating hospitals for self-reflection and action to improve IPC practices by the health care workers. Secondly, the findings will be disseminated to the University of Dodoma for academic purposes. Third, the findings will be shared with the Ministry of Health policy and guideline reviews; and lastly, the findings will be disseminated to international conferences and publication in peer reviewed journals for international audience.

## Discussion

Evidence is available from previous studies to indicate that exposure to practices of HCWs’ that do not adhere to principles of IPC can lead to puerperal sepsis or neonatal sepsis[4,10,24,25]. Nevertheless, puerperal sepsis and neonatal sepsis are stand still problems in the Sub Saharan Africa that take countless lives of maternal women and neonates[2]. The level of health care facility, say regional referral hospital, district hospital and/or health centre and either government owned facility or private facility drive or hinder adherence to IPC standards as influenced by shortage or lack of supplies, inadequate training and inadequate organizational support for safe practice[19,26]. This study first assesses HCWs’ pattern of practice in adhering to IPC standards and uses a longitudinal approach to trace the path of women from entry to the antenatal/labor ward until discharge with her neonate and examine the points where exposure to infection is possible. The finding of this study will help to identify infection preventable practices and their magnitude of risk for puerperal and neonatal sepsis in the context of labor and postnatal wards and inform the possible and relevant interventions.

## Data Availability

This is a protocol paper, data are not yet available.

## Ethics statement

The University of Dodoma Research Ethics Committee (UDOM-REC) approved this study with Ref No. MA.84/261/’A’/25. All participants recruited signs informed consent before participation in this study.

## Public Involvement Statement

It was not appropriate or possible to involve patients or the public in the design of our research protocol. Thus, pubic involvement was not implemented.

## Consent for publication

Not applicable.

## Author’s contribution

**L.E.M** conceived the study, developed the study plan/protocol and supervises acquisition of, will do data analysis and draft the manuscript. **G.M** conceptualized and shaped the study idea, reviewed the study plan/protocol, reviewed the research tools provides intellectual technical guidance of the study.

## Availability of data

Data will be available from the corresponding author upon request after completion of collection and analysis.

## Acknowledgements

The research authors would like to appreciate support from the Ministry of Health Tanzania and the University of Dodoma for their partial financial support to this study.

## References

1 Tomczyk S, Storr J, Kilpatrick C, et al. Infection prevention and control (IPC) implementation in low-resource settings: a qualitative analysis. Antimicrob Resist Infect Control 2021;10:1–11. doi:10.1186/s13756-021-00962-3

2 World Health Organisation. AIDE-MEMOIRE GLOBAL ALERT AND RESPONSE Core components of infection prevention and control programmes in health care Background. 2011;:2.

3 WHO. WHO recommendations for prevention and treatment of maternal peripartum infections. 2015. https://www.who.int/publications/i/item/9789241549363

4 Buddeberg BS, Aveling W. Puerperal sepsis in the 21st centuryC: progress, new challenges and the situation worldwide. 2015;:1–7. doi:10.1136/postgradmedj-2015-133475

5 MoHCDGEC. National Infection Prevention and Control Standards for Hospitals in Tanzania. Dar es Salaam, Tanzania: 2020.

6 Arianpoor A, Zarifian A, Askari E, et al. “Infection prevention and control idea challenge” contestC: a fresh view on medical education and problem solving. 2020;:1–10.

7 World Health Organization. Infection prevention and control Guidance to action tools. 2021.

8 Friday O, Edoja O, Osasu A, et al. Assessment of infection control practices in maternity units in Southern Nigeria. 2012;24:634–40.

9 Ngonzi J, Bebell LM, Fajardo Y, et al. Incidence of postpartum infection, outcomes and associated risk factors at Mbarara regional referral hospital in Uganda. 2018;:1–11.

10 C. Kajeguka D, Reuben Mrema N, Mawazo A, et al. Factors and Causes of Puerperal Sepsis in Kilimanjaro, Tanzania: A Descriptive Study among Postnatal Women who Attended Kilimanjaro Christian Medical Centre. East African Heal Res J 2020;4:158–62. doi:10.24248/eahrj.v4i2.639

11 Meharun-Nissa Khaskheli, Shahla Baloch AS. Risk factors and complications of puerperal sepsis at a tertiary healthcare centre. 2013;29:972–6.

12 Demisse GA, Sifer SD, Kedir B, et al. Determinants of puerperal sepsis among post partum women at public hospitals in west SHOA zone Oromia regional STATE, Ethiopia (institution BASEDCASE control study). 2019;:1–6.

13 Tirivavi E, Chikanya V, Mundagowa PT. Infection Control Practices Associated with Puerperal Sepsis in Harare City Infection Control Practices Associated with Puerperal Sepsis in Harare City Maternity Units. Published Online First: 2019. doi:10.11648/j.cajph.20190501.15

14 Johnson-Walker YJ, Kaneene JB. Epidemiology: Science as a Tool to Inform One Health Policy. Beyond One Heal From Recognit to Results 2018;:3–30. doi:10.1002/9781119194521.ch1

15 Satyarup D, Kumar M, Dalai RP, et al. Theories of disease causation: An overview. Indian J Forensic Med Toxicol 2020;14:8075–9. doi:10.37506/ijfmt.v14i4.12923

16 Dicker RC, Coronado F, Koo D, et al. Lesson 1: Introduction to Epidemiology. In: Principles of Epidemiology in Public Health Practice, Third Edition. An Introduction to Applied Epidemiology and Biostatistics. 2012. 1–52. https://www.cdc.gov/csels/dsepd/ss1978/ss1978.pdf

17 Mor G, Cardenas I. The Immune System in Pregnancy: A Unique Complexity. Am J Reprod Immunol 2010;63:425–33. doi:10.1111/j.1600-0897.2010.00836.x

18 Basha S, Surendran N, Pichichero M. Immune responses in neonates. Expert Rev Clin Immunol 2014;10:1171–84. doi:10.1586/1744666X.2014.942288

19 Kinyenje E, Hokororo J, Eliakimu E, et al. Status of Infection Prevention and Control in Tanzanian Primary Health Care Facilities: Learning From Star Rating Assessment. Infect Prev Pract 2020;2:100071. doi:10.1016/j.infpip.2020.100071

20 Song JW, Chung KC. Observational studies: Cohort and case-control studies. Plast Reconstr Surg 2010;126:2234–42. doi:10.1097/PRS.0b013e3181f44abc

21 Mann JC. Observational research methods. Research design II: cohort, cross sectional, and case-control studies. Emerg Med J 2003;20:54–60.http://search.ebscohost.com/login.aspx?direct=true&db=eoah&AN=6675644&site=ehost-live

22 NBS. Tanzania in Figures 2018 National Bureau of Statistics Dodoma. 2019.

23 Gage AD, Yahya T, Kruk ME, et al. Assessment of health facility quality improvements, united republic of Tanzania. Bull World Health Organ 2020;98:849–858A. doi:10.2471/BLT.20.258145

24 Cabal A, Schmid D, Lepuschitz S, et al. Nosocomial outbreak of Streptococcus pyogenes puerperal sepsis. Clin Microbiol Infect 2019;25:521–3. doi:10.1016/j.cmi.2018.11.028

25 Borghesi A, Mazzucchelli I, Pozzi M, et al. IS-013 Neonatal Sepsis, New Preventive Strategies. Arch Dis Child 2014;99:A5.1-A5. doi:10.1136/archdischild-2014-307384.13

26 Akagbo SE, Nortey P, Ackumey MM. Knowledge of standard precautions and barriers to compliance among healthcare workers in the Lower Manya Krobo District, Ghana. BMC Res Notes 2017;:1–9. doi:10.1186/s13104-017-2748-9

